# Exploring the Attitudes, Barriers, Concerns, Differences, and Ethics (ABCDEs) of Healthcare Decision Making of Vagus Nerve Stimulation

**DOI:** 10.64898/2026.10.27.26366559

**Authors:** Christi RP Sullivan, Logan R Caola, Rachel A Johnson, Cash Differding, Anita Randolph, Angie Meija, Danniella Balangoy, Alik S Widge, Thomas R Henry, Ziad Nahas, Laura Y Cabrera, Saydra R Wilson

**Affiliations:** University of Minnesota, Department of Psychiatry and Behavioral Health Sciences, Minneapolis, MN; Just the Pill/Abortion Delivered, Minneapolis, MN; Masonic Institute for the Developing Brain, University of Minnesota, Minneapolis, MN; University of Minnesota, Department of Neurology, Minneapolis, MN; The Pennsylvania State University, Department of Engineering Science and Mechanics, Center for Neural Engineering, University Park, PA, The Pennsylvania State University, Rock Ethics Institute, University Park, PA. The Pennsylvania State University, Huck Institutes of the Life Sciences, University Park, PA

**Keywords:** Vagus Nerve Stimulation (VNS), Treatment-Resistant Depression (TRD), Epilepsy, Health Care Access, Medical Stigma Informed, Consent

## Abstract

Vagus nerve stimulation (VNS) is an established neuromodulation therapy approved for treatment-resistant depression (TRD) and epilepsy, with growing interest in its application to other conditions. Despite demonstrated clinical efficacy, VNS remains underused in psychiatric care. This mixed-methods study examined stakeholder perceptions of VNS, focusing on access, stigma, ethical concerns, and acceptability across epilepsy, depression, heart failure (HF), and rheumatoid arthritis (RA). We conducted six focus groups and 20 semi-structured interviews with patients, clinicians, and one support person, and integrated qualitative themes with quantitative data on stigma and treatment barriers. Participants revealed stark disparities in VNS adoption: epilepsy patients reported fewer insurance barriers, while TRD participants faced delayed approvals, high out-of-pocket costs, and limited access to specialty care. Depression stakeholders reported higher stigma scores and expressed greater reluctance to consider surgical treatment, despite possessing comparable knowledge to epilepsy stakeholders. Ethical concerns—particularly regarding informed consent and impaired decision-making—surfaced frequently in the context of TRD. Most participants agreed that the appropriateness of VNS should align with disease severity and impact on quality of life. Many cautiously supported VNS for HF and RA, provided strong efficacy data supports its use. Hope emerged as a consistent theme, especially among TRD participants seeking alternatives after exhausting other treatments. These findings underscore the need to address insurance barriers, reduce stigma, and enhance provider education. As VNS expands into new indications, policymakers and clinicians must implement reforms that promote equitable access, decentralized care, and ethically sound consent practices to ensure inclusive neuromodulation therapy adoption.

## Introduction

### History of VNS

Cervical vagus nerve stimulation (VNS) is an established neuromodulation therapy that involves implanting a small pulse generator in the chest to deliver electrical impulses to the left (typically) vagus nerve. The vagus nerve is mixed with a preponderance of sensory fibers. It connects various organs to the brainstem, with outputs into the parasympathetic system and somatic sensory system and plays a crucial role in modulating immune responses through the cholinergic anti-inflammatory pathway (Mandalaneni & Rayi, 2025). By modulating vagal activity, VNS has shown efficacy in reducing seizures in epilepsy and alleviating symptoms of treatment-resistant depression (TRD). Despite nearly three decades of clinical use, questions persist regarding the role of stigma, ethical concerns, and barriers to access in the uptake of VNS across various mental health conditions. VNS received U.S. Food and Drug Administration (FDA) approval for epilepsy in 1997 and for Major Depressive Disorder (MDD) in 2005. Its mechanism of action involves altering neuronal excitability through sustained vagal modulation, likely involving polysynaptic actions at multiple brain sites (Vonck & Larsen, 2018). Over 125,000 devices have been implanted for epilepsy and approximately 5,000 for depression, underscoring a notable discrepancy (“Implantable Vagus Nerve Stimulators Market to Reach $1.27 Billion in 2033, Forecasts GlobalData,” 2023). While it offers long-term symptom relief, its adoption in psychiatric conditions is relatively limited, highlighting potential barriers beyond clinical efficacy (Aaronson et al., 2022; O’Reardon et al., 2006)

### Characteristics of VNS

VNS is unique in the field of neurostimulation. Unlike deep brain stimulation (DBS), which penetrates the cranium, VNS surgery is minimally invasive because the surgery is shorter, quicker, and does not involve the brain. Additionally, while transcranial magnetic stimulation (TMS) generally involves multiple weeks of on-site sessions, VNS offers a more prolonged and sustained course of treatment. Once implanted, a VNS battery can last anywhere from five to 15 years (*Vagus Nerve Stimulation (VNS)*, n.d.). VNS has several advantages, including the less invasive nature of surgery, and demonstrated long-term efficacy, particularly in reducing seizure frequency in epilepsy patients and improving symptoms of treatment-resistant depression, sometimes outperforming pharmaceutical interventions (Aaronson et al., 2017; Batson et al., 2022). Though common side effects include voice alterations, difficulty swallowing, and shortness of breath, treatment is generally regarded as safe. Side effects resulting from programming can often be reduced through changes in stimulation parameters. Many patients experience delayed but meaningful benefits, with seizure or depressive symptom relief often becoming apparent over months to years (Aaronson et al., 2022; O’Reardon et al., 2006). Approximately 60% of epilepsy patients and 40-60% of TRD patients respond to VNS within 2 years (Aaronson et al., 2022; Englot et al., 2016; O’Reardon et al., 2006).

### Stigma considerations

VNS sits at the intersection of treatments for both “physical” (epilepsy) and “mental” (depression) conditions, both of which carry distinct forms of stigma (Kwon et al., 2022; Pescosolido et al., 2021; Schnyder et al., 2017). While epilepsy has historically been shrouded in myths of demonic possession, depression remains burdened by the misconception that individuals are personally responsible for their condition, highlighting how stigma manifests in distinct yet equally harmful ways for both disorders. Stigma is both an attitudinal barrier and an ethical concern that significantly impacts medical technology uptake. Mental illness stigma can decrease treatment-seeking and exacerbate social isolation. Similarly, stigma can lead to fear and feeling of rejection that cause lower levels of treatment seeking in epilepsy which has been linked to lower quality of life and increased seizure frequency (*Addressing Stigma Surrounding Epilepsy | Epilepsy Foundation*, n.d.; Kwon et al., 2022; Mayor et al., 2022). The progress made in reducing stigma around epilepsy may partially account for the lag in adoption of VNS within the mental health field.

### Ethical considerations

Ethical questions around informed consent and patient autonomy arise for both depression and epilepsy with some individuals presenting with impaired decision-making capacity. Treatments commonly used to treat these diseases can improve aspects of cognition relevant to decision making for some, but others can experience short- or long-term impairments to decision-making capacity (Goering et al., 2017, 2021; Hendriks et al., 2019; Jotterand et al., 2010; Kwan & Brodie, 2001; Novak et al., 2022; Rock et al., 2014). The need for adequate informed consent processes has been a pillar of medical ethics to date. In addition to treatment decision-making, adequately informing patients of the delayed effects is a central topic. Access concerns relating to travel and resource limitations also raise justice issues: VNS is typically offered at academic medical centers, leaving patients in rural areas or states lacking such centers at a disadvantage (*States with the Most Academic Medical Centers*, n.d.). Given that VNS requires frequent in-person visits during the first year to fine-tune the device, exploring how to address barriers to make VNS more accessible is a priority.

### Stakeholder considerations

Healthcare decision-making is a two-way process that involves clinicians and patients. As gatekeepers of this implantable neuromodulation therapy, clinicians significantly influence patient access through their knowledge, biases, and willingness to present or refer to VNS as an option. Meanwhile, patients with limited exposure to new VNS research may harbor misconceptions or concerns regarding invasiveness and safety, further shaping decisions (Cabrera et al., 2023).

### Study rationale

Despite VNS’s clinical longevity, the interplay of stigma, ethical concerns, and limited accessibility remains understudied. As VNS expands to treat conditions such as rheumatoid arthritis, heart failure, fibromyalgia and other chronic medical conditions, (Lerman et al., 2025), a deeper examination of stakeholder perceptions will guide ethical, practical, and policy-related discussions, ultimately informing better adoption and informed consent practices. Of particular interest is how stigma does or does not develop in the context of disease about which one does not have significant personal knowledge. This study aims to understand stakeholders’ perceptions of VNS through reports of healthcare barriers, ethical considerations, perceived acceptability, and knowledge level.

## Methods

### Methodology

We used a mixed methods approach to capture broad (questionnaire) and in-depth (focus group and interview) insights into stakeholder perceptions of VNS. The Phase 1 focus groups informed refinements to the Phase 2 semi-structured interviews. Participants were recruited via purposive sampling to ensure representation across disease types (Depression or Epilepsy) and roles (Patient, Clinician, and Support Person). This study was approved by the [REDACTED FOR REVIEW] Institutional Review Board (STUDY00020128), and all participants provided informed consent before data collection.

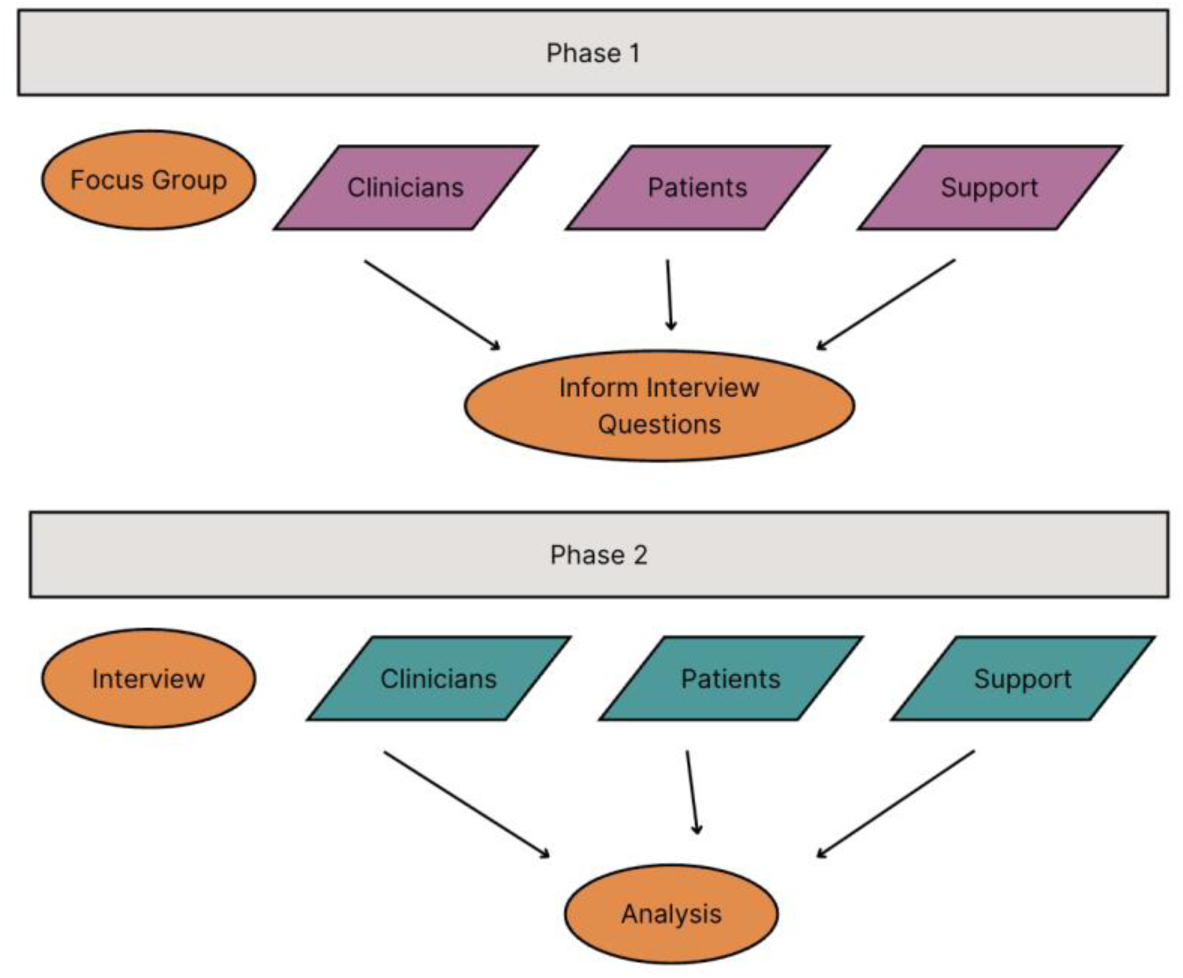

### Recruitment and Sampling

Patients were recruited from the University of Minnesota Physicians St. Louis Park clinic, which treats TRD and Epilepsy patients. Initially, we targeted individuals in the parent REVEAL study pipeline [1U54AT012307-01]. REVEAL aims to understand the impact of VNS on various organ systems such as cardiovascular, metabolic, immune, and gastrointestinal functions. Due to slow accrual, we later included non-REVEAL epilepsy patients. Clinicians who delivered VNS for depression or epilepsy were recruited through the LivaNova physician directory and referrals from REVEAL study clinicians. We identified support persons by asking participants to recommend a caregiver or family member for the study.

### Inclusion and Exclusion Criteria

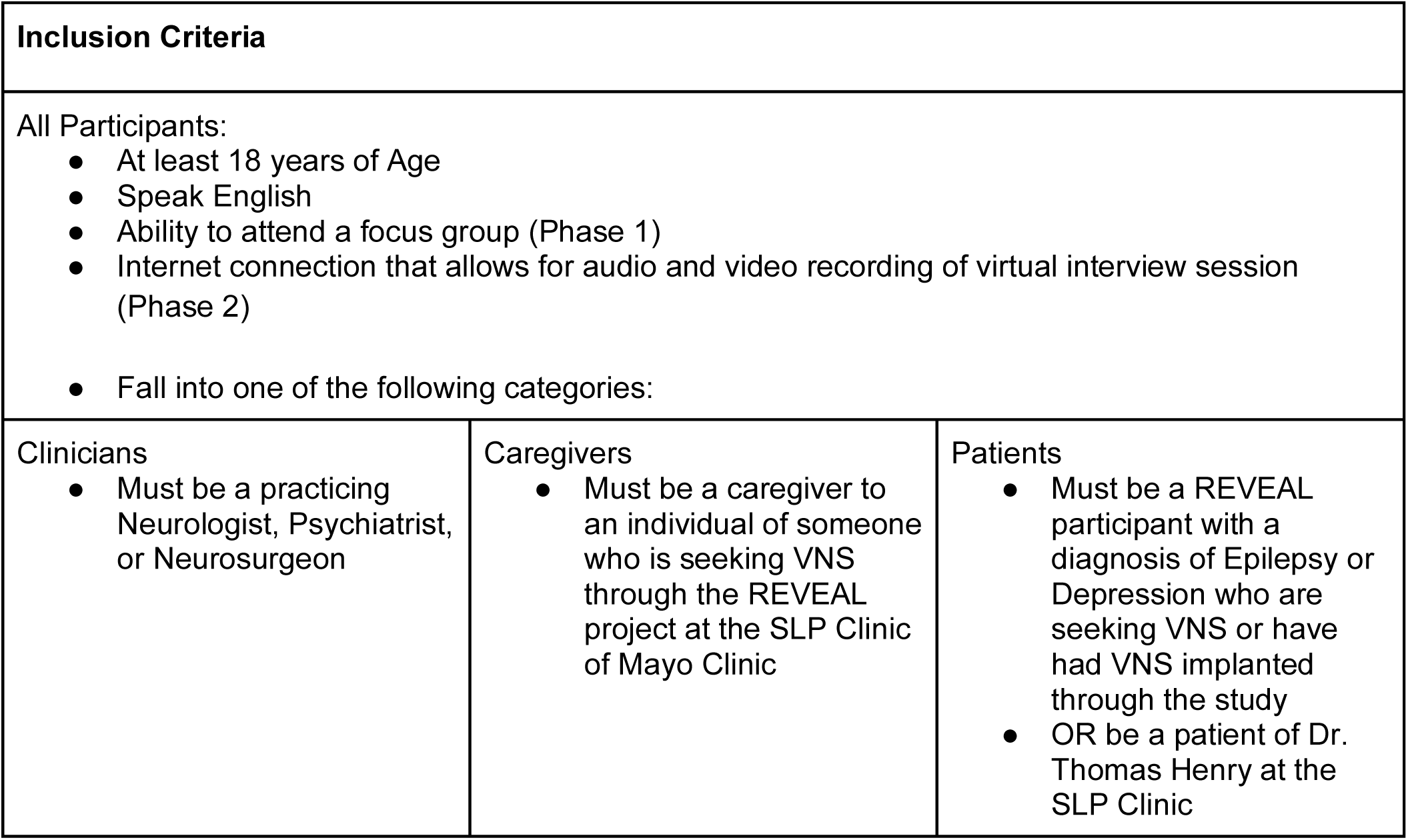
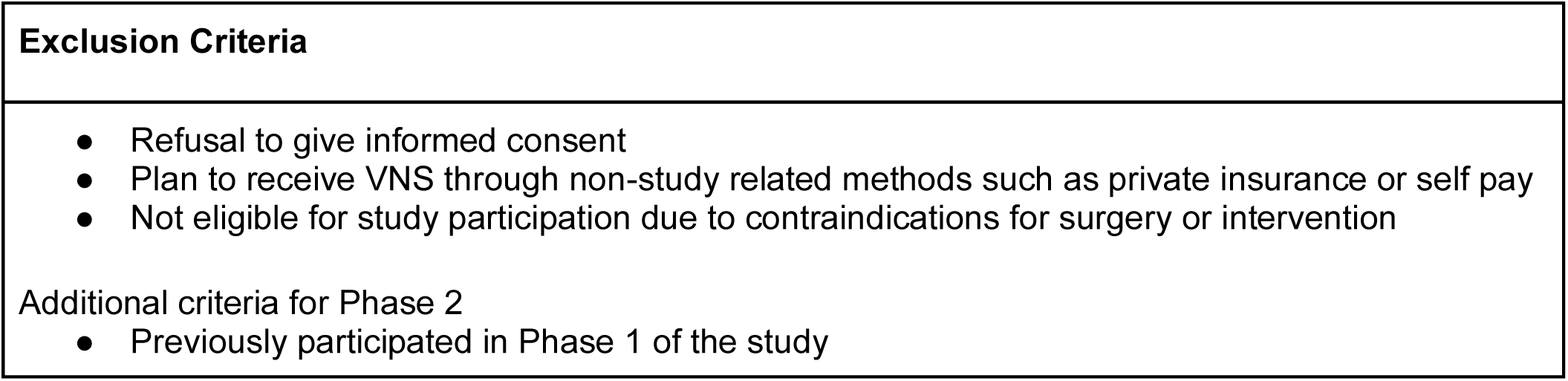

#### Phase 1

Six individuals participated in focus groups organized by stakeholder groups. Before each focus group, participants completed online questionnaires measuring VNS knowledge, perceived barriers, attitudes toward VNS, and stigma using the 8-item Stigma Scale for Chronic Illnesses (SSCI-8). We conducted two 90-minute focus groups between March and April 2024. One, with three depression patients, took place in person on the University of Minnesota campus, while the other, with two depression clinicians and one epilepsy clinician, was held via Zoom. Focus groups for epilepsy patients, epilepsy caregivers, and depression caregivers were planned, but recruitment efforts were unsuccessful. Both sessions were facilitated by Anita Randolph, PhD, using a semi-structured guide and then transcribed. These discussions generated preliminary insights that shaped Phase 2 interview questions and informed the development of a customized semi-structured interview template for each stakeholder group.

#### Phase 2

We recruited 21 new participants to complete in-depth, semi-structured interviews alongside the same questionnaires from Phase 1. Each received vignettes on a secondary condition (Heart Failure (HF) or Rheumatoid Arthritis (RA)) to facilitate questionnaire completion. A study team member conducted interviews via secure Zoom, focusing on VNS invasiveness, barriers to care, stigma, comparative treatment preferences, and attitudes toward using VNS for a hypothetical secondary condition. All sessions were recorded and transcribed using Zoom’s transcription feature.

### Data Analysis

We conducted a data-driven thematic analysis using Dedoose (Braun & and Clarke, 2006). Two independent coders used the interview guide to inform an initial codebook. As interviews were conducted, new codes were added to reflect emergent concepts. After at least six interviews, data collection continued until two consecutive interviews yielded no new codes, which was our operational definition of thematic saturation. At that point the coders refined the codebook by merging overlapping codes and reclassifying codes that had few occurrences, generally less than 5 occurrences in the set of interviews, into broader thematic categories when appropriate.

Each transcript was first coded by the person who conducted the interview using the original codebook, then re-coded by the non-interviewing coder using the revised codebook to reduce interviewer bias. Both coders jointly coded three transcripts (approximately 15% of the dataset) to assess agreement and refine coding decisions; fewer than three discrepancies per transcript emerged during this process, all of which were resolved through discussion and consensus.

Themes were initially developed independently by each coder and then consolidated collaboratively. The final thematic structure was organized into overarching domains that reflected stakeholder perceptions of VNS. Questionnaire data were exported to RStudio for descriptive and inferential statistical analysis (e.g., t-tests) and integrated into Dedoose to allow for mixed-methods comparisons.

Finally, qualitative themes were compared with quantitative findings to generate a comprehensive understanding of stakeholder perspectives.

### Participant Characteristics

A total of 92 individuals were approached to participate in this study. Of those, 30 met the inclusion criteria and consented to participate, of which 26 participated in either a focus group or a semi-structured interview. Figure 1 breaks down the recruitment and enrollment process via a CONSORT diagram.

**Figure 1:**
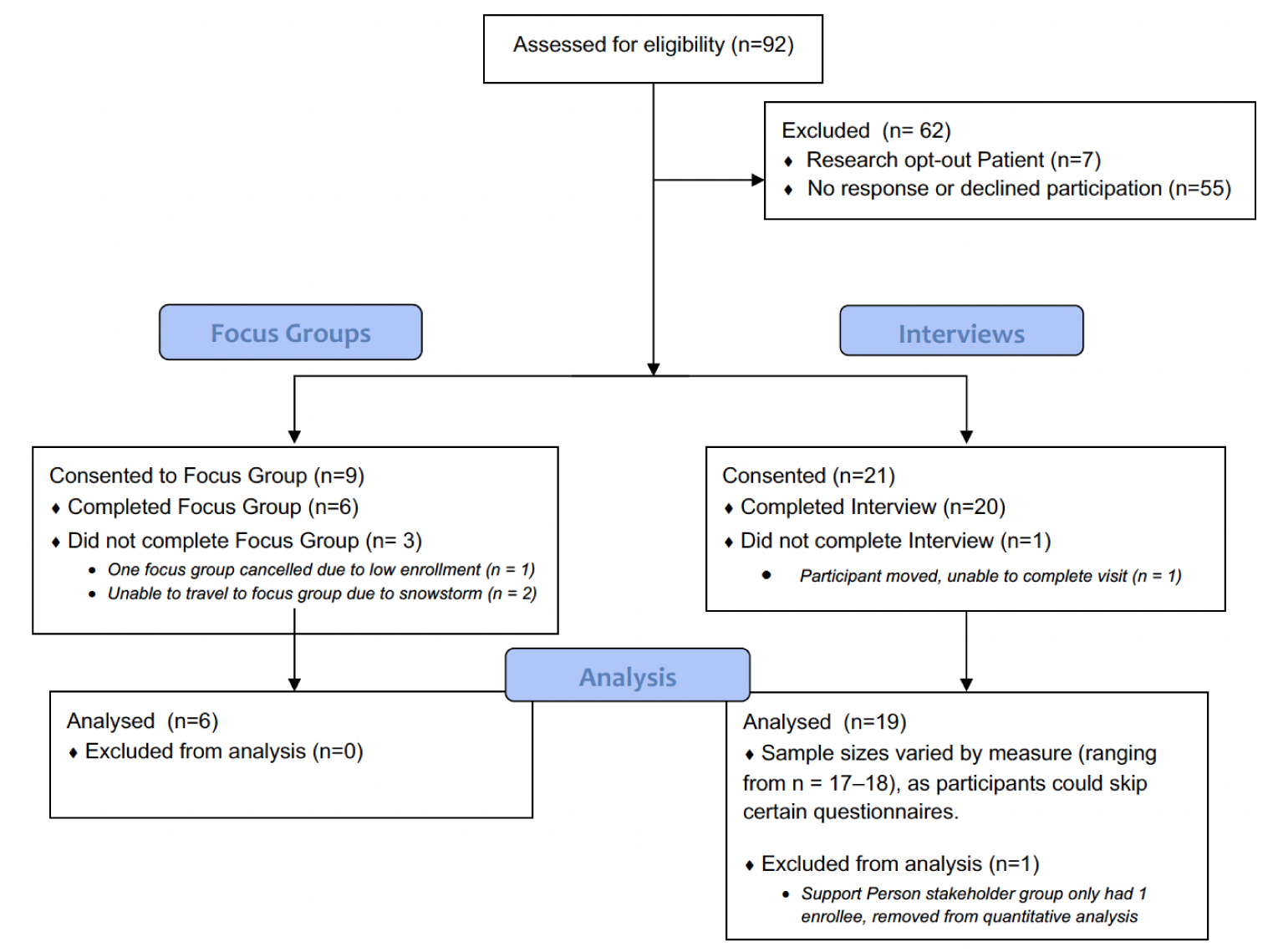
CONSORT diagram

**Table 1.**
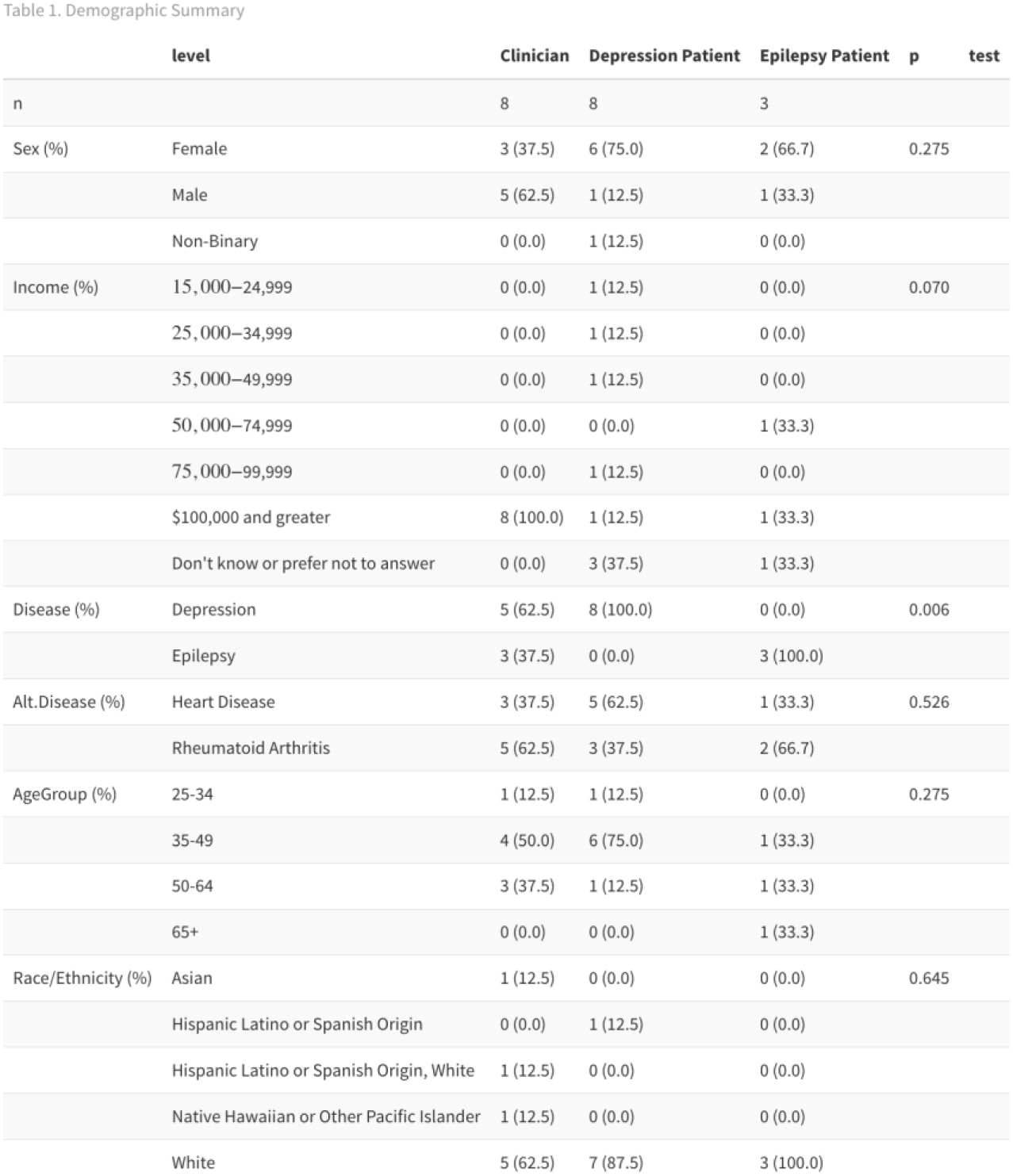
summarizes the demographic characteristics of the final sample.

**Phase 1 (Focus Groups).** Six individuals participated in one of two focus groups: a group of three Depression patients and a mixed group of three Clinicians (two Depression Clinicians and one Epilepsy Clinician). **Phase 2 (Semi-Structured Interviews).** A new set of 20 individuals completed semi-structured interviews, including eight Depression patients, three Epilepsy patients, eight Clinicians, and one Support Person. At the same time, this study aimed to include three distinct stakeholder groups (Clinicians, Patients, and Support Persons). The final sample included only one Support Person, as slow recruitment and low availability in the Depression cohort limited participation. Consequently, analyses on support-person perspectives were not included in this analysis.

In Phase 2, the mean participant age was 44.9 years (SD = 11.2). Overall, 58% identified as Female, 37% as Male, and 5% as Non-Binary. Among the 19 interview participants in the final analysis, 70% were Depression stakeholders, and 30% were Epilepsy stakeholders. The mean duration of illness was 27 years (SD = 3.74) for Depression and 5 years (SD = 4.58) for Epilepsy. All eight Depression patients had tried medication, therapy, and light therapy; 63% (n=5) had undergone ECT, 50% (n=4) had tried TMS, 37% (n=3) had utilized residential treatment, and 25% (n=2) had tried Ketamine. The eight clinicians practiced in diverse settings, with 50% working in Academic Health Centers, 25% in Private Practice, 12.5% in Tertiary Care Centers, and 12.5% in Community Practices.

## Results

In the following sections, we present our findings on (1) how VNS acceptability and access differ by disease group (epilepsy vs. depression), (2) how clinicians’ and patients’ perspectives converge or diverge, and (3) whether and why participants would support VNS for new conditions (e.g., rheumatoid arthritis, heart disease). **The focus of the result section is on interview data from Phase 2.** We integrate quantitative results from Phase 2 (e.g., stigma scales, barrier frequencies) and qualitative insights (focus group/interview quotes) to illustrate these findings. Quotes have been cleaned of typos and other utterances for clarity and fluidity.

### Thematic Findings

#### Access and Insurance

One of the most prominent themes was insurance coverage and overall access to VNS. Seventeen discussed access as a key issue, and sixteen of the nineteen interview participants mentioned insurance issues at least once. Depression participants more frequently ranked the cost of treatment as their top barrier, while epilepsy participants focused on side effects or disease minimization. An analysis of overall barrier rankings (p = 0.003) revealed that Clinicians and Patients differ in prioritizing obstacles to receiving VNS. Figure 2 overviews the difference in average ranking of treatment barriers by the disease group.

**Figure 2.**
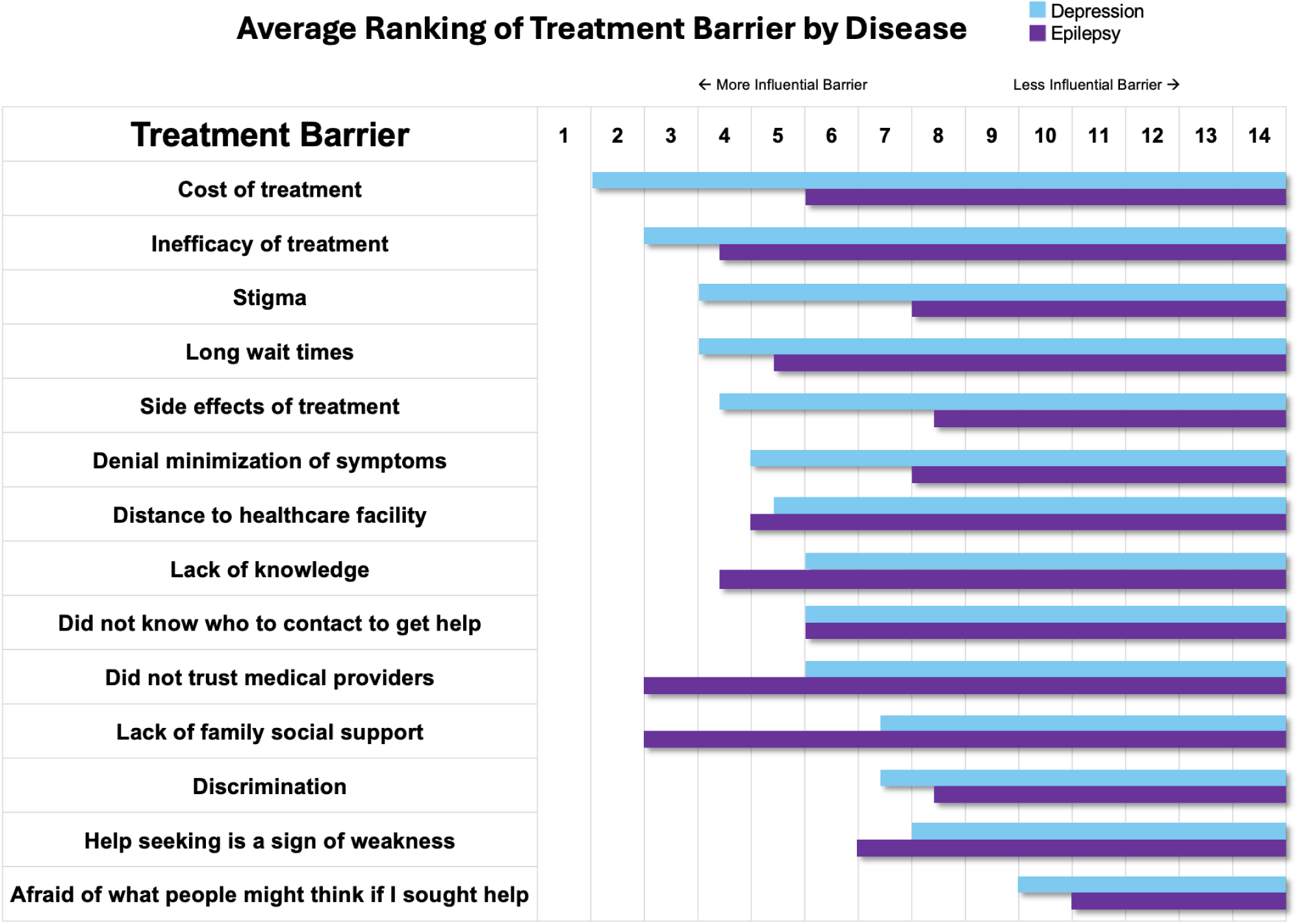
Average ranking of treatment barriers, broken down by disease, sorted by most to least influential by Depression.

This overall difference is categorized by the barriers brought up by clinicians compared to patients. Clinicians focused on insurance coverage as a key aspect of access:

> *“[VNS] still remains … a very niche set up … depending upon [the] ivory tower academic centers.” – Depression Clinician*
>
> *“[There are] a ton of barriers… Insurance [companies] are very reluctant to pay $150,000 for implanting a VNS. They don’t really see the benefit.” – Depression Clinician*

Clinicians also discussed other key aspects affecting access to care, including transportation, cost, and time off work, and how these are intermixed with social determinants of health:

> *“There [are] many people in the United States that … don’t have access to proper care, whether it be because of money, … race, or both, [or] because of areas of where they live.” – Epilepsy Patient*

Epilepsy clinicians found coverage of VNS for their indication easy to obtain, attributing ease of access partly to the high costs of emergency care for uncontrolled seizure:

> *“My experience is I send a patient for consultation for VNS, and within 2–3 weeks they’re in for … the consultation, and then another couple of weeks the VNS is implanted.” – Epilepsy Clinician*
>
> *“[Center for Medicaid Services] is not reimbursing for VNS [for Depression] … while it is routinely covered by insurance for Epilepsy.” – Epilepsy Clinician*
>
> *“If they keep on seizing, they go to the ED… it’s cheaper for them to have [VNS].” – Epilepsy Clinician*

In contrast, Depression clinicians noted that long insurance wait times added additional access barriers for their patient population:

> *“[Barriers are] transportation, cost, time off work, childcare, insurance coverage.” – Depression Patient*
>
> *“There’s a long duration of waiting [for insurance approval]. But then, is the patient even still willing to do [the] procedure if we get them covered? … Difficulty with insurance, and duration of time to approval is a main limiting factor for patients.” – Depression Clinician*
>
> *“VNS is something you have to plan, get consent for, discuss with the patient, convince the patient. It takes a few months just to get the patient convinced and starting the process. And then you need the surgical consultation and then you need a surgical date for OR … It’s more of a long term plan for someone who had multiple, multiple episodes and relapses … but is not so acutely ill that they cannot provide consent or make a decision.” – Depression Clinician*

Patients noted the complex relationship between insurance coverage and physical access to care:

> *“I don’t think any insurance will cover it as of right now. And I saw that … it was anywhere between $12,000 to $18,000. And that’s just for the surgery itself.” – Depression Patient*

Depression clinicians and patients described long delays in receiving VNS approval, often months of waiting or needing research studies to obtain VNS for TRD.

> *“My health insurance wouldn’t cover VNS. So this is … a once in a lifetime opportunity to be able to participate in this study and to get to. I have this device implanted without having to pay a billion dollars to try this.” – Depression Patient*

In sum, insurance disparities, limited access to the treatment, and out of pocket costs pose significant obstacles to VNS adoption across all conditions, particularly for Depression and any emerging indications.

#### Stigma

Stigma emerged as a critical theme, with quantitative scores showing Depression participants reporting significantly higher experiences of blame or unkindness and scoring higher across stigma questions. In particular, they were substantially more likely to agree with the statement “Some people acted as though it was my fault I have this illness” (p = 0.001) and “Because of my illness people were unkind to me” (p=0.008) after adjusting for multiple comparisons. In total, 13 of 19 participants brought up themes of stigma, spanning all subgroups.

> *“I am very vocal and open about my mental health struggles, but… people are taught that it’s such a shameful thing to have mental health… [it’s] not your fault.” – Depression Patient*

Both patients and clinicians noted a cultural bias where psychiatric conditions are sometimes minimized. One Depression participant explained:

> *“I feel like other people might judge you more if you say we’re doing it for depression… I think some people for depression would probably be like, ‘What are you so sad about that you need an implant?’” – Depression Support Person*
>
> *“Hey, I have epilepsy, and I have this device implanted, there’s not really cultural stigma around that. Now, if I say I have depression, I have a device implanted. People are like, “What are you doing?” So there’s still stigma around mental health issues, and especially something as big as a surgery.” – Depression Clinician*

Depression patients and their clinicians often felt Depression was not taken as seriously as Epilepsy and worried about being judged for considering an implantable device:

> *“I mean, the epilepsy… it can really be dangerous. Depression, I have a hard time seeing that the same as a physical condition… just ‘Oh, pull yourself up by your bootstraps.’” – Depression Patient*
>
> *“People take epilepsy more seriously; it seems more real.” – Depression Clinician*

Epilepsy patients noted stigma, too, but from a more “medical/physical illness” lens. They sensed less pushback on having a device implanted for a recognized neurological disorder. Clinicians echoed patients’ observations regarding mental health conditions being viewed as “less legitimate” or “willed away”:

> *“Some think of depression as a choice… that it should be easy to treat if you do psychotherapy or yoga…” – Depression Clinician*

Participants illustrated the internal and external stigmas associated with seeking a surgical treatment for depression: ”:

> *“You know, it’s easier if a patient with refractory epilepsy comes to you [for VNS]. But [Depression] patients that have had hundreds of ECT [treatments] and you suddenly say VNS and they say ‘whoa, whoa, whoa, I’m not **that** depressed. What are you talking about? No, no, no surgery, you’re crazy.’” – Depression Clinician*
>
> *“I don’t think that depression really needs any devices put into your body … I think it’s more of you having to deal with it yourself.” – Epilepsy Patient*

Interestingly, a participant’s skepticism about VNS for arthritis underscores how new or emerging indications may also confront doubts and stigma about whether a condition is deemed “worthy” of implantable interventions.

> *“I have chronic pain that I deal with and … another reason for participating in [REVEAL] is to see what happens with [pain] as a secondary thing. And when I was talking with the surgeon, he was like “Chronic pain is off the table. Get it out of your head that that’s going to improve”. So that’s why I don’t think that [VNS is appropriate for] arthritis.” – Depression Patient*

However, any surgical implant for “invisible” conditions, including RA and HF, could still face residual skepticism, although participants largely believed mental health carried the greatest stigma burden.

#### Invasiveness and Treatment Order

Nearly all participants acknowledged that VNS is surgically invasive, yet less invasive than penetrating brain surgeries (e.g., DBS, cingulotomy). Both clinicians and patients made the comparison of VNS to a pacemaker, citing its placement in the chest rather than the skull.

> *“It’s certainly invasive… but pacemakers are invasive too.” – Depression Patient*
>
> *“I use the analogy to a pacemaker because most people kind of understand that that goes under the skin and is there in the chest wall.” – Epilepsy Clinician*
>
> *“It doesn’t seem terribly invasive… sometimes I think drugs are more invasive.” – Epilepsy Patient*
>
> *“[Regarding brain surgery] That’s like messing with the mainframe, and you just don’t mess with the mainframe.” – Depression Patient*

Epilepsy participants often embraced earlier VNS use if seizures were uncontrolled. In contrast, Depression participants noted it “happens very late” in their treatment journey, especially if standard therapies (medications, therapy, TMS, ECT) fail. In our sample the individuals had lived with Depression for an average of 27 years, compared to Epilepsy with an average of 5 years.:

> *“VNS happens very late in the [Depression] patient journey… it’s less likely to be as helpful as it is for epilepsy.” – Depression Clinician*
>
> *“If I tried 2 or 3 medications …and they’re not really seizure free, then we have that conversation [of pursuing surgery].” – Epilepsy Clinician*

Participants also stressed a treatment order, with VNS typically introduced after several failed medication trials. There was a general belief that *“the level of the treatment must match the level of disease” – Depression Clinician*, meaning that the invasiveness of VNS must be relational to the disease it is treating. Participants believed earlier VNS could be advantageous if the disease was irreversible (e.g., RA joint damage), but still felt less-invasive options should be tried first:

> *“You can’t turn back the clock on RA, you can only slow it down.” – Depression Patient*
>
> *“It shouldn’t be the first mode of approach… but it’s the insurance company that decides that.” – Depression Patient*
>
> *“If medications aren’t working… VNS seems like a worthwhile thing to try.” – Depression Patient*

VNS is also potentially more manageable for families and support persons to accept compared to an invasive brain surgery like DBS or a cingulotomy:

> *“My experience is most caregivers don’t want [their loved ones] to have brain surgery because they are disabled. And it’s really hard to do the work-up. So that’s why we end up using the VNS mostly.” – Epilepsy Clinician*
>
> *“If they benefit from epilepsy surgery, we go with [that]… if not, then we go with… VNS, because it’s been around the longest.” – Epilepsy Clinician*

Clinicians emphasized physical surgical risk:

> *“If you go by invasiveness… medication is less invasive than VNS, because it’s not surgery.” – Epilepsy Clinician*
>
> *“It’s an implanted device… You can have some risk of infections with the surgery, but then it works.”* – *Depression Clinician*

Patients had a diverse view of invasiveness of VNS compared to other treatments, mentioning that medications and therapy carry their own types of invasiveness. Patients saw VNS as invasive, but sometimes saw ECT as more invasive given personal experience with long term side effects

> *“Some medications can have long lasting impacts … I think most medications wouldn’t be as invasive, but maybe some with longer term impacts would be.” – Depression Patient*
>
> *“VNS is more physically invasive, but therapy uncovers emotional things that can’t be undone…”* – *Depression Patient*
>
> *“My only parlor trick is my memory… with ECT, that’s a big thing you’re probably losing.” – Depression Patient*

Moreover, no one saw VNS as a standalone therapy; they expected it to be paired with medications, therapy, or other adjuncts.

> *“It’s sort of like they’re complementary… you need multiple modalities.” – Depression Clinician*

#### Quality of Life and Appropriateness of VNS

Of the 19 interviewees, nine were asked to consider HF, and 10 were asked to consider RA. Quality of life (QOL) consistently emerged as a key driver of whether stakeholders would consider VNS. Participants across Depression, Epilepsy, HF, and RA unanimously agreed that a severely diminished QOL could justify the surgical risk:

> *“Even if it doesn’t [help everything], maybe it would help with my mood, maybe it would help with the pain.” – Depression Patient*
>
> *“Living with VNS sounds less dangerous than living with a heart condition.” – Depression Patient*
>
> *“If they’re not responding to medication, quality of life is poor… I think we should give them a chance with another treatment.” – Epilepsy Clinician*
>
> *“Especially if quality of life is being affected. I think you reach a certain point where it’s worth trying out almost anything … While VNS is invasive there are risks that [are] not super significant or likely.” – Depression Patient*

There were no significant differences between clinicians and patients in their overall ratings of VNS appropriateness for Depression and Epilepsy. However, Epilepsy clinicians tended to rate VNS as slightly less appropriate for Depression. This difference points to the potential for clinicians to stigmatize treatments. Despite similar conclusions, each group arrived at them in different ways as seen in Figure 3.

**Figure 3:**
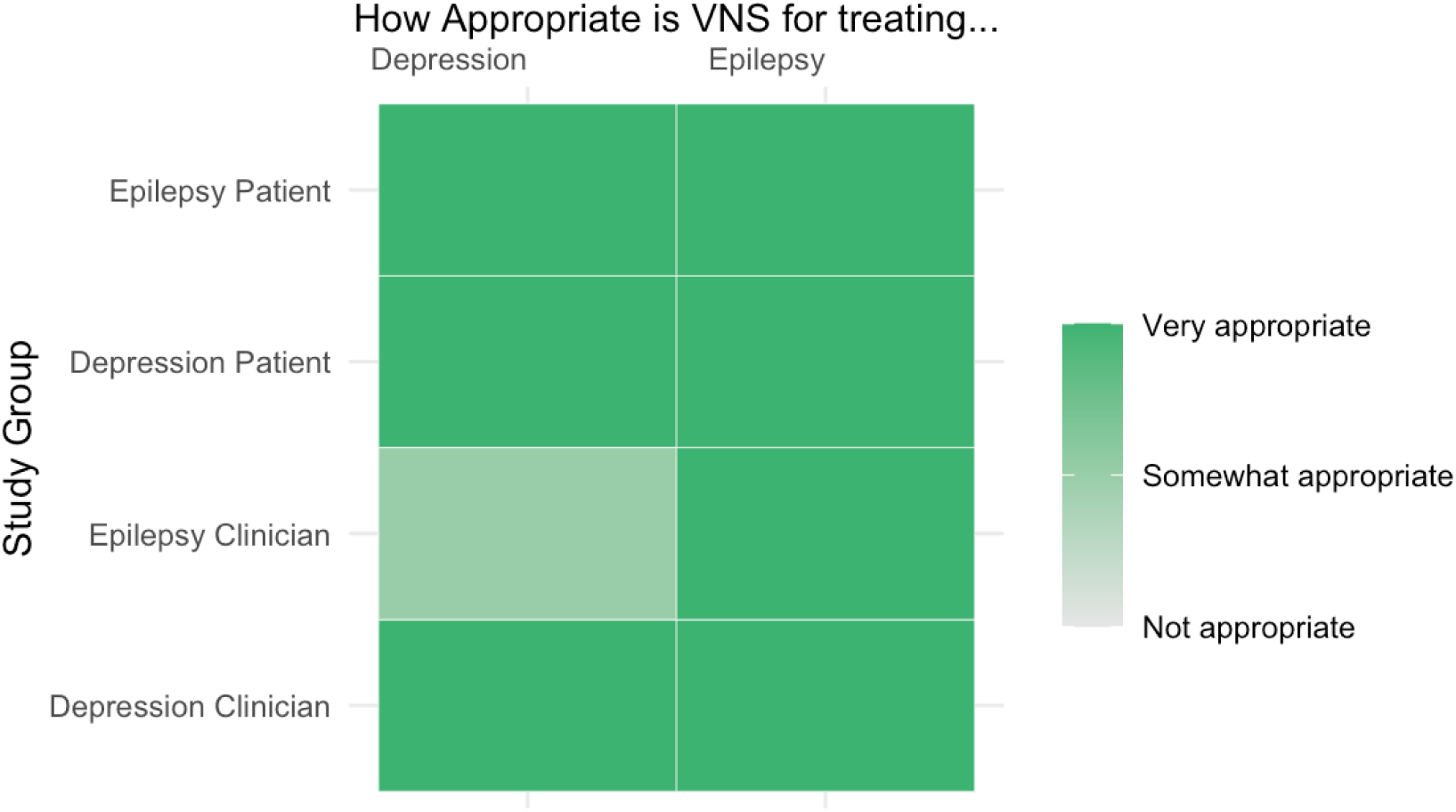
Stakeholder views of appropriateness for using VNS for Depression and Epilepsy

Figure 4 shows stakeholder ratings of VNS appropriateness for HF/RA. Epilepsy clinicians were somewhat open to using VNS for these conditions, whereas Depression clinicians wanted stronger efficacy data before adopting VNS. Patients, particularly those with Epilepsy, were surprisingly receptive if QOL was significantly impaired:

> *“Epilepsy seems more cut and dry… so I assume it’s probably more effective.” – Depression Patient*
>
> *“If it could help anyone with either [Epilepsy or Depression]… I would certainly consider it.” – Epilepsy Patient*

Most stakeholders generally expressed tentative acceptance of new VNS indications if quality of life was severely compromised, underscoring the common refrain that “the level of treatment must match the level of disease.”

Most participants anticipated less stigma for RA or HF than for Depression, likening HF to well-accepted cardiac implants and RA to a physical pain condition:

> *“We’re used to having… implants for heart issues.” – Depression Patient*
>
> *“I don’t think people with RA would care as much… it’s just another medical treatment.” – Depression Clinician*
>
> *“I mean, it has to do with your physical quality of life … your heart is going out feels like that’s … more pressing to me than you feel sad all the time.” – Depression Patient*

Both clinicians and patients worried that Heart Failure or Rheumatoid Arthritis (RA), which are still experimental for VNS, might face even higher barriers:

> *“Some of these new medications are really good, so I’m not sure what room there is for VNS in RA.” – Clinician*
>
> *“Insurance coverage… what if that changed? … having to get the battery replaced was another thing… the insurance coverage and [future] costs.” – Depression Patient*
>
> *“[…]for other conditions, it probably will boil down to the cost. Will it be cheaper?” – Epilepsy Clinician*

#### Ethical Concerns

Participants unanimously agreed that VNS should match the level of disease severity. For treatment-resistant Depression or uncontrolled Epilepsy, it might be justified; otherwise, a less-invasive approach should be prioritized.

> *“With depression, you are implanting in people who are considered treatment resistant… Are you taking advantage of someone’s desperation?” – Depression Clinician*
>
> *“There’s such a wide range of how severe you have epilepsy or depression that you’d have to make your decision off of the severity of those before going ahead with it.” – Epilepsy Patient*

Capacity to Consent surfaced repeatedly (14/19 interviews), especially for severe Depression, where cognition or desperation might impair fully informed decisions.

> *“VNS is something you have to plan, get consent for, discuss with the patient, convince the patient… It’s more of a long-term plan…” – Depression Clinician*
>
> *“Hopefully … if they truly can’t consent they have a person or a proxy that can do it [for them].” – Depression Patient*
>
> *“I think making patients very aware of any risks involved, financial costs and the emotional risks*
>
> *… making sure that patients have the informed consent that they are not going in blindly.” – Depression Patient*

Clinicians consistently requested robust clinical evidence, especially for new indications:

> *“I don’t really know the data for depression… I’m familiar with [VNS] data for epilepsy.” – Epilepsy Clinician*
>
> *“We need evidence… I won’t be surprised if it’s similar efficacy, but I’d need to see the data.” – Depression Clinician*

Overall, while patients often based their views on perceived disease severity and personal empathy, clinicians underscored the scientific rationale for whether VNS should be offered in the first place.

#### Hope

Despite financial and logistical hurdles, a strong undercurrent of hope emerged throughout the interviews. Many living with longstanding or treatment-resistant conditions expressed optimism for at least partial relief:

> *“VNS is giving me a little more hope that I can live like this [waves hand], instead of this [nosedive motion]… I would almost do anything to not live that way.” – Depression Patient*
>
> *“And when I heard about it being used for depression, I was like, wow, that - that would really be cool.” – Depression Patient*
>
> *“They’ve gotten to this point … that they’ve exhausted from trying so many different things and kind of. Not getting the results that they need to …live a life worth living …This is another hopeful option.” – Depression Patient*

However, for some Depression participants these hopes were guarded, shaped by prior disappointments with ECT, TMS, or multiple medications:

> *“I don’t put a lot of hope into thinking, ‘This is the one’… if it works, it works, and if it doesn’t, it doesn’t.” – Depression Patient*
>
> *“So, that’s kind of my mindset is, it’s not so much hopelessness, but it’s also not putting all your eggs in one basket, you know.” – Depression Patient*

Epilepsy participants valued the possibility of fewer seizures and better daily functioning without the same feeling of guarded hope.

> *“If it were a case where … someone was really struggling with it … on a daily basis. Then, yes, I mean [I’d] definitely want to have [VNS].” – Epilepsy Patient*

Patients described seeking therapy that could improve their life, in a way making “life worth living,” while clinicians embraced the notion of hope but emphasized keeping realistic expectations due to the gradual time-to-response. Hope extended to HF/RA, where some participants reasoned that if standard approaches fail, VNS might “offer another hopeful option,” especially when QOL is severely compromised.

## Discussion

Collectively, these interwoven themes reveal how stakeholders navigate VNS across multiple conditions. While Depression and Epilepsy remain the primary contexts for approved VNS, participants indicated cautious openness toward HF/RA if robust data supports its efficacy and the disease severely impairs daily functioning. Overall, the tension between financial/insurance barriers, mental health stigma, and the potential for symptom relief shaped stakeholder views on whether and how VNS should be adopted. These findings set the stage for a deeper discussion on the implications for clinical practice, policy, and future research.

Our study highlights how stakeholders perceive and adopt VNS across Epilepsy, Depression, and potential new indications (HF/RA). While participants generally recognized VNS’s therapeutic value, they also voiced concerns about financial barriers, directly attributed to insurance coverage policies and delayed coverage approval, as well as social stigma, particularly regarding mental health. At the same time, a strong theme of hope prevailed, with many participants viewing VNS as an opportunity for significant improvement in QOL, providing a robust body of evidence supporting its use.

Our findings are consistent with literature suggesting that neuromodulation for psychiatric conditions lags behind more “physical” indications due to greater stigma (Cabrera et al., 2023). In our sample the stigma associated with VNS is rooted in the disease it is treating instead of the treatment itself. Individuals were generally open to VNS as a treatment but their opinions changed based on the disease it was treating. Knowledge of VNS did not differ between the stakeholder groups, reinforcing the notion that differences in attitudes towards VNS is not due to a knowledge deficit. For instance, although Epilepsy also carries some stigma, Depression participants faced higher levels of shame, blame, and hesitancy to undergo an implantable intervention. Within our sample it was unclear whether this stigma was self stigma, externalized stigma, or a mix of the two. This disparity mirrors the broader pattern of device adoption seen with deep brain stimulation (DBS): around 300,000 patients have received DBS implants for Parkinson’s disease, while only 300 implants have been performed for obsessive-compulsive disorder (Mocking et al., 2021). Such stark differences reflect how mental health stigma can hinder acceptance and coverage of surgical neuromodulation, and even the funding of research for different diagnoses, despite comparable clinical promise.

The role of insurance coverage emerged as a powerful determinant of VNS adoption. Patients who lacked coverage, often Depression patients, reported waiting months or relying on research studies to access VNS, underscoring the need for cost-effectiveness data to persuade insurers. These results extend prior observations that lack of coverage disproportionately hinders the adoption of invasive therapies for mental health compared to epilepsy or movement disorders (Mocking et al., 2021). Policymakers and insurers might benefit from systematically evaluating the long-term cost savings of VNS, particularly for TRD and emerging conditions.

Going forward, researchers must collect robust, high-quality data that demonstrate VNS’s safety, efficacy, and cost-benefit advantage for new indications. Such evidence could help streamline insurance approval pathways, reducing out-of-pocket costs, and broadening patient access.

Additionally, efficacy and cost-benefit must be viewed in relation to the disease. As our population attested, Epilepsy is easier to get approval for because the outcome is quantitative; You can observe a countable reduction in seizure numbers from VNS. With Depression, untreated or poorly managed depression is associated with increased risk for substance abuse, chronic conditions, such as cardiovascular disease, and premature mortality (Remes et al., 2021); However, it is harder to get at these endpoints because TRD trials are often focused on changes in subjective clinical ratings of patient symptoms. A prime example of this in practice is the RECOVER trial of VNS for TRD. In this trial, VNS for TRD was assessed for QOL benefits through 6 outcome surveys and was deemed to be superior to no stimulation, but this was only true for three of the six surveys (Rush et al., 2024). For outcomes that are less objective, it is harder to identify and prove efficacy of a treatment.

Understanding the unique stigma of a disease is especially crucial for HF and RA, where VNS remains experimental. It is unclear how either disease group would receive VNS as a treatment and what stigma may emerge, and how it will impact uptake. We know that treatment disparities can be rooted in the disease it is treating. For example, the comparatively lower uptake of VNS for Depression mimics the historical pattern seen in DBS for OCD vs. Parkinson’s Disease. Clinicians, advocacy groups, and policy initiatives should jointly tackle mental health stigma, ensuring that implantable interventions are not dismissed as “too invasive” for psychiatric disorders, and proactively consider what role stigma may play in VNS for future indications. Educational efforts targeting clinicians and patients, highlighting the physical underpinnings of TRD and the efficacy of VNS, may erode the biases currently dissuading many from considering this option.

Participants consistently stressed that “the level of treatment must match the level of disease.” Clarifying where VNS fits the treatment trajectory, especially for new indications, would help integrate it into standard practice. Integrating VNS earlier for irreversible or highly debilitating conditions might prevent years of suffering, provided patients are fully informed and clinically appropriate.

Ethical concerns about the capacity to consent were especially salient for severely depressed individuals. Formal assessments of decision-making capacity and structured consent protocols can minimize the risk of inadvertently taking advantage of patient desperation. However, the due diligence of minimizing risk should be balanced with the time and effort required of potential surgical candidates. As shown by our study population, individuals considering VNS have often lived with their disease for years, if not decades, having tried numerous other treatments. The consent process should match the capacity to consent of the individual instead of lumping all individuals into a group of high vulnerability. Considerations for how current treatment regimens may be impacting capacity to consent should be explored, acknowledging that fluctuation in consent ability can be influenced by both the disease progression and the medications being used to treat it. Educating patients and potential proxies on the long timeline to VNS response is likewise essential for realistic expectations and adherence. Multiple types of neuromodulation (TMS, DBS, VNS) have some level of delayed response, making the emphasis on time to response central to consent discussions.

Travel distance and reliance on academic centers limit uptake of Neuromodulation treatments, including VNS. Exploring decentralized models such as telemedicine follow-ups, remote programming, training of residents to expand clinician access, or closer collaboration with community clinics may help expand access. An example of this model is the Abbott NeuroSphere Virtual Clinic for neurostimulation devices which allows for routine device checks, programing adjustments, and initiation of new settings through a real time visit done via video call (*NeuroSphere^TM^ Digital Care*, n.d.) Early data shows that remote care can be consistently and reliably delivered through remote care platforms (Silburn et al., 2022). Providing special training for non-academic clinicians to manage and program VNS would similarly reduce the burden on patients living far from major medical centers.

### Limitations

Several factors constrain the generalizability of our findings. First, only one support person ultimately participated, limiting our ability to draw robust conclusions about caregivers’ perspectives. This was in part due to low enrollment in the Epilepsy arm and the lack of identified support persons from our Depression participants. The reliance on recruitment from a parent study inherently limited the pace of enrollment. For example, in December 2024 when recruitment for this study was ending, there were 26 depression enrollments in contrast to 2 epilepsy enrollments. Additionally, in asking Depression patients to identify a support person of theirs who they could recommend to the study, most people replied that they felt they didn’t have anyone who provided significant support. It is unclear if this is due to stigma surrounding the disease, lack of support experienced by people living with depression, social isolation, or other factors.

In our case, relying on a neuroethics supplement is what allowed us to get this study funded. Limited flexibility in the timing of supplements can create vulnerabilities. If the parent study faces delayed IRB approval, study stoppage due to audits, or slow/low enrollment, it can be difficult for the neuroethics supplement to adjust with limited funds and timing.

Snowy Minnesota winter weather further complicated focus group enrollment and attendance, reducing the number of focus groups we were able to run and limiting participation to only 3 persons per group, which is smaller than normal for focus groups. Our sample may be biased toward individuals already somewhat familiar or open to VNS, especially those in the parent research pipeline. Working within the single site of the parent study also inherently limited the diversity of perspectives given the homogeneity of our clinic sample. Lastly, while we collected insights on HF/RA, these were hypothetical vignettes; actual experiences might differ once VNS is formally tested or approved for those conditions.

## Conclusion

To address disparities in VNS uptake within Depression and Epilepsy and reduce disparities for future indications, large, multi-site research is needed to explore VNS’s adoption in diverse socioeconomic and geographic settings. Emphasizing telehealth integration to minimize travel burdens and conducting a cost-benefit analysis comparing the long-term costs of treating conditions with standard treatments versus VNS could be particularly impactful. This type of analysis can help determine when VNS becomes a financially viable option for insurance approval. Early focus on robust data and cost-effectiveness may ease the path to insurance coverage and bolster confidence among clinicians, payers, and patients.

VNS is widely recognized by clinicians and patients in our sample for its potential to improve quality of life. Yet, it remains underutilized for mental health indications due to stigma, insurance barriers, and ethical concerns regarding the capacity to consent. As VNS expands to new indications like HF and RA, these challenges must be addressed proactively through rigorous efficacy studies, educational campaigns to reduce stigma, and policies promoting equitable access to realize the therapy’s benefit fully. Our findings illuminate the complex interplay between clinical, social, and financial factors that shape whether and how VNS is adopted, offering a foundation for more inclusive and evidence-informed approaches to neuromodulation.

## Data Availability

All data produced in the present study are available upon reasonable request to the authors.

## Statements and Declarations

Financial interests: The authors declare they have no financial interests.

## Funding

**Neuroethics Supplement** - 3U54AT012307-01S1 Exploring the Attitudes, Barriers, Concerns, Differences, and Ethics (ABCDEs) of Healthcare Decision Making in Prospective Peripheral Indications for Vagus Nerve Stimulation Patients

## Parent Grant

1U54AT012307-01 REVEAL - Research Evaluating Vagal Excitation and Anatomical Linkages

The study team would like to acknowledge the collaborating PIs of the parent study for their leadership and foundational work that opened the avenue for this supplement to come to fruition: Dr. John Osborn, PhD; Dr. Lynn E. Eberly, PhD; Dr. Sayeed Ikramuddin, MD, MHA; Dr. Hubert H. Lim, PhD; Dr. Vaughan Macefield, PhD; and Dr. Catherine A. McCarty, PhD.

The University of Minnesota provides REDCap to researchers using Grant Number UM1TR004405 from the National Institutes of Health’s National Center for Advancing Translational Sciences

LC would like to thank the support of her Dorothy Foehr Huck and J. Lloyd Huck Chair in Neuroethics.

Supported in part by the Masonic Institute for the Developing Brain, University of Minnesota.

Informed consent was obtained from all individual participants included in the study.

## Notes

### Competing Interest Statement

The authors have declared no competing interest.

### Author Declarations

IRB of University of Minnesota gave ethical approval for this work.

