## Supplemental file for "Exploring the Attitudes, Barriers, Concerns, Differences, and Ethics (ABCDEs) of Healthcare Decision Making of Vagus Nerve Stimulation"

### Appendix A

**Goal of Focus Groups** = Determine themes we want to drill into deeper in interviews, identify questions that clearly have a wide range of responses/content that needs to be explored and eliminate questions that have clear consensus from interviews
***90 minutes of interview time***

**Goal of Interviews** = Collect individual experiences to form a larger narrative. Ask more complex questions. Get assessment of stigma, invasiveness, seek treatment, etc. Put the invasiveness/perception of VNS on the scale of all neuromodulation.
***60 minutes of interview time***

#### Schedule of Events

| | ***Focus Group*** | ***Interview*** |
|---|---|---|
| ***Pre-Session*** | ● Demographic Survey<br>● Knowledge Survey<br>● Attitudes Survey<br>● Stigma Questionnaire<br>● Rankings<br>● Barriers<br>● Support Activities (Caregiver only) | ● Demographic Survey<br>● Knowledge Survey<br>● Attitudes Survey<br>● Stigma Questionnaire<br>● Rankings<br>● Barriers<br>● Support Activities (Caregiver only) |
| ***During Session*** | ● Questions | ● Questions |

##### *Focus Group Questions*

Physicians (Depression and Epilepsy Single Group)

1. GENERAL (20 minutes)
    1. What sort of practice are you a part of (community, hospital, academic, etc.)?
    2. What role do you play in the delivery of VNS?
    3. How effective do you feel VNS is for [DEPRESSION/EPILEPSY]? Is VNS more, less, or similarly effective as other treatments for [DEPRESSION/EPILEPSY]?
    4. What do you think are the benefits of VNS compared to psychotherapy or taking pharmaceuticals? [note similarities and differences]
    5. What do you think are the risks of VNS compared to psychotherapy or taking pharmaceuticals? [note similarities and differences]

6. Do you think VNS for [DEPRESSION/EPILEPSY] is more beneficial than for [EPILEPSY/DEPRESSION]?
2. HEALTHCARE DECISION MAKING (30 minutes)
    1. When in treatment trajectory do you suggest VNS? When/How do you start to introduce the option of VNS?
    2. What role does insurance coverage play in your decision to/not to recommend VNS?
    3. What should patients be focused on when deciding about VNS therapy?
    4. What is the primary concern that patients voice about VNS?
    5. Would you feel differently if this was VNS for a different indication [DISEASES]?
3. INVASIVENESS (15 minutes)
    1. How invasive do you perceive VNS? Why?
    2. Do you think VNS is more or less invasive than undergoing psychotherapy? Why?
    3. Do you think VNS is more or less invasive than taking pharmaceuticals? Why?
    4. Do you think VNS is more or less invasive than excisional surgery/cingulotomies? Why?
    5. Do you think VNS is more or less invasive than diet changes? Why?
4. BARRIERS (15 minutes)
    1. What are the main barriers you see patients encounter when trying to access VNS?
    2. What sort of post surgery follow-up/support would be ideal?
5. WRAP UP (10 minutes)

***Focus Group Questions***

Patient (Depression)

1. GENERAL (20 minutes)
    1. How effective do you think current treatments for depression are?
    2. Briefly, what types of depression treatments have you tried so far?
    3. Briefly, how effective do you think it/they have been?
    4. Before your provider recommended VNS, what did you know about it?
    5. Why did you decide to try VNS?
    6. How effective do you feel VNS is for your indication? How effective are other treatments?
    7. How do you anticipate VNS will compare to other therapies you have tried?
2. HEALTHCARE DECISION MAKING (30 minutes)
    1. What do you think are the benefits of VNS compared to psychotherapy or taking pharmaceuticals? [note similarities and differences]
    2. What do you think are the risks of VNS compared to psychotherapy or taking pharmaceuticals? [note similarities and differences]
    3. Do you think VNS for DEPRESSION is more beneficial than for EPILEPSY? [Please explain why]
    4. Are the risks and side effects worth the potential benefits of VNS?
    5. Would your opinions about the benefits and risk change if it was being used to treat other indications, like rheumatoid arthritis or heart failure? Why? What changes?
3. INVASIVENESS (15 minutes)
    1. How would you describe how invasive VNS is?  Why?  In what sense?
    2. Do you think VNS is more or less invasive than undergoing psychotherapy? Why?

    3. Do you think VNS is more or less invasive than taking pharmaceuticals? Why?
4. BARRIERS (15 minutes)
    1. Do you have any concerns about barriers to accessing VNS?
    2. Are there things that you think might keep people from being able to use VNS? (e.g., distance, waiting times, side effects, transportation)?
    3. Is the slow onset of symptom relief a barrier?
    4. What sort of post surgery follow-up/support would be ideal?
5. WRAP UP (10 minutes)

***Focus Group Questions***

Patient (Epilepsy)

1. GENERAL (20 minutes)
    1. How effective do you think current treatments for epilepsy are?
    2. Briefly, what types of epilepsy treatments have you tried so far?
    3. Briefly, how effective do you think it/they have been?
    4. Before your provider recommended VNS, what did you know about it?
    5. Why did you decide to try VNS?
    6. How effective do you feel VNS is for your indication? How effective are other treatments?
    7. How do you anticipate VNS will compare to other therapies you have tried?
2. HEALTHCARE DECISION MAKING (30 minutes)
    1. What do you think are the benefits of VNS compared to taking pharmaceuticals? [note similarities and differences]
    2. What do you think are the risks of VNS compared to psychotherapy or taking pharmaceuticals? [note similarities and differences]
    3. Do you think VNS for EPILEPSY is more beneficial than for DEPRESSION?
    4. Are the risks and side effects worth the potential benefits of VNS?
    5. Would your opinions about the benefits and risk change if it was being used to treat other indications, like rheumatoid arthritis or heart failure? Why? What changes?
3. INVASIVENESS (15 minutes)
    1. How would you describe how invasive VNS is? Why? In what sense?
    2. Do you think VNS is more or less invasive than excisional surgery/cingulotomies? Why?
    3. Do you think VNS is more or less invasive than taking pharmaceuticals? Why?
    4. Do you think VNS is more or less invasive than diet changes? Why?
4. BARRIERS (15 minutes)
    1. Do you have any concerns about barriers to accessing VNS?
    2. Are there things that you think might keep people from being able to use VNS? (e.g., distance, waiting times, side effects, transportation)?
    3. Is the slow onset of symptom relief a barrier?
    4. What sort of post surgery follow-up/support would be ideal?
5. WRAP UP (10 minutes)

***Focus Group Questions***

Caregiver (Depression)

1. GENERAL (20 minutes)
    1. How effective do you think current treatments for depression are?
    2. How did you react when your loved one decided to have surgery? [stigma]
    3. Before discussing VNS with a provider, what did you know about it?
    4. How effective do you feel VNS is for Depression?
    5. How do you anticipate VNS will compare to other therapies your loved one has tried?
2. HEALTHCARE DECISION MAKING (30 minutes)
    1. What do you think are the benefits of VNS compared to psychotherapy or taking pharmaceuticals? [note similarities and differences]
    2. What do you think are the risks of VNS compared to psychotherapy or taking pharmaceuticals? [note similarities and differences]
    3. Do you think VNS for DEPRESSIONis more beneficial than for EPILEPSY?
    4. Are the risks and side effects worth the potential benefits of VNS?
    5. Would your opinions about the benefits and risk change if it was being used to treat other indications, like rheumatoid arthritis or heart failure? Why? What changes?
3. INVASIVENESS (15 minutes)
    1. How would you describe how invasive VNS is?  Why?  In what sense?
    2. Do you think VNS is more or less invasive than undergoing psychotherapy? Why?
    3. Do you think VNS is more or less invasive than taking pharmaceuticals? Why?
4. BARRIERS (15 minutes)
    1. Do you have any concerns about barriers to accessing VNS?
    2. Are there things that you think might keep people from being able to use VNS? (e.g., distance, waiting times, side effects, transportation)?
    3. Is the slow onset of symptom relief a barrier?
    4. What sort of post surgery follow-up/support would be ideal?
5. WRAP UP (10 minutes)

***Focus Group Questions***

Caregiver (Epilepsy)

1. GENERAL (20 minutes)
    1. How effective do you think current treatments for epilepsy are?
    2. How did you react when your loved one decided to have surgery? [stigma]
    3. Before discussing VNS with a provider, what did you know about it?
    4. How effective do you feel VNS is for Epilepsy? How effective are other treatments?
    5. How do you anticipate VNS will compare to other therapies your loved one has tried?
2. HEALTHCARE DECISION MAKING (30 minutes)
    1. What do you think are the benefits of VNS compared to diet changes/resection surgery or taking pharmaceuticals? [note similarities and differences]
    2. What do you think are the risks of VNS compared to diet changes/resection surgery or taking pharmaceuticals? [note similarities and differences]

3. Do you think VNS for EPILEPSY is more beneficial than for DEPRESSION?
4. Are the risks and side effects worth the potential benefits of VNS?
5. Would your opinions about the benefits and risk change if it was being used to treat other indications, like rheumatoid arthritis or heart failure? Why? What changes?
3. INVASIVENESS (15 minutes)
    1. How would you describe how invasive VNS is?  Why?  In what sense?
    2. Do you think VNS is more or less invasive than undergoing resection surgery? Why?
    3. Do you think VNS is more or less invasive than taking pharmaceuticals? Why?
4. BARRIERS (15 minutes)
    1. Do you have any concerns about barriers to accessing VNS?
    2. Are there things that you think might keep people from being able to use VNS? (e.g., distance, waiting times, side effects, transportation)?
    3. Is the slow onset of symptom relief a barrier?
    4. What sort of post surgery follow-up/support would be ideal?
5. WRAP UP (10 minutes)

***Interview Questions***

Physicians (Depression and Epilepsy Single Group)

1. GENERAL (10 minutes)
    1. What sort of practice are you a part of (community, hospital, academic, etc.)?
    2. What role do you play in the delivery of VNS?
        1. Who else is involved in the decision to screen eligibility of patients for VNS?
    3. How effective do you feel VNS is for [DEPRESSION/EPILEPSY]? Is VNS more, less, or similarly effective as other treatments for [DEPRESSION/EPILEPSY]?
    4. What do you think are the benefits of VNS compared to psychotherapy or taking pharmaceuticals? [note similarities and differences]
        i. Prompt as needed: Confers resilience to stress and adversities (when it works) Less adverse effects [compared to medications]
    5. What do you think are the risks of VNS compared to psychotherapy or taking pharmaceuticals? [note similarities and differences]
    6. Do you think VNS for [DEPRESSION/EPILEPSY] is more beneficial than for [EPILEPSY/DEPRESSION]?
    7. Do patients typically come to you asking for VNS or are you the one introducing VNS for the first time?
        i. How do you inform patients about VNS (e.g. industry video, access to patient group with VNS, other)
        ii. When do you inform patients about VNS?
2. RISKS (15 minutes)
    1. Is VNS more, less, or about the same risk level as [TREATMENT]
    2. Does VNS have more, less, or about the same amount of benefit as [TREATMENT]
    3. Does VNS have more, less, or about the same amount of side effects as [TREATMENT]
    4. Are the risks and side effects worth the potential benefits of VNS? What are the side effects expected from VNS? Compared to [TREATMENT] do patients have any control over the side effects of VNS?

        i. Prompt as needed: cannot have imaging from torso
    1. Would you feel the same about your patients seeking VNS for other indications, like [rheumatoid arthritis or heart failure/other]?
3. INVASIVENESS (15 minutes)
    1. How would you describe how invasive VNS is?  Why?  In what sense?
    2. Do you think VNS is more or less invasive than undergoing psychotherapy?Why?  How so?
    3. Do you think VNS is more or less invasive than taking pharmaceuticals?  Why?  How so?
    4. Do you think VNS is more or less invasive than excision surgery or Deep Brain Stimulation?  Why?  How so?
        1. Prompt as needed: Does the fact that the surgery does not enter the skull influence your view on invasiveness?
    5. Again, considering other diseases such as [Epilepsy, Depression, Rheumatoid Arthritis, Heart failure] does the invasiveness of VNS seem appropriate for the disease it would be treating?
4. BARRIERS (15 minutes)
    1. Do you have any concerns about practical barriers to accessing these therapies, if patients want to use them? Are there things that you think might keep people from being able to use these treatments? (e.g., distance, waiting times, side effects, transportation, surgery, understanding, lack of education medical community)?
    2. What do you see as the most important ethical questions we should answer about using VNS for treating [depression/epilepsy/ OTHER]  Why?  How so?
        1. Prompt as needed: lack of equity/who gets access. When and for whom it will work. Delayed response
    c. Do you think the barriers to VNS depend on the indication?
    d. When you think of the word ‘support’ in relation to VNS what comes to mind?
    Prompt: What would make people feel more supported?
    Prompt: Any specific concerns about demographics such as race, age, sex, culture, religion, etc.
5. WRAP UP (5 minutes)

***Interview Questions***

Patient (Depression)

1. GENERAL
    1. Briefly, how long have you lived with Depression? How many types of treatments have you tried?
    2. How does VNS compare to other therapies you have tried?
    3. VNS is used for both epilepsy and depression. From your understanding of the disease, do you think VNS for epilepsy is more, less, or about the same level of benefit compared to depression?
2. RISK
    1. Is VNS more, less, or about the same risk level as [TREATMENT]
        i. Prompt as needed: slow onset of VNS
    1. Does VNS have more, less, or about the same amount of benefit as [TREATMENT]

        1. Prompt as needed: about expectations regarding medication reduction or potential for less side effects
    2. Does VNS have more, less, or about the same amount of side effects as [TREATMENT]
    3. Are the risks and side effects worth the potential benefits of VNS? What are the side effects expected from VNS? Compared to [TREATMENT] do you have any control over the side effects of VNS?
        1. Prompt as needed: about surgical complications, perceived inability to do certain imaging procedures
    4. If you had another disease such as [rheumatoid arthritis or heart failure] and were offered VNS as a treatment would you consider it? Why or why not?
3. INVASIVENESS
    1. How would you describe how invasive VNS is? Why? In what sense?
    2. Do you think VNS is more or less invasive than undergoing psychotherapy? Why? How so?
    3. Do you think VNS is more or less invasive than taking pharmaceuticals? Why? How so?
    4. Do you think VNS is more or less invasive than Deep Brain Stimulation? Why? How so?
        1. Prompt as needed: Does the fact that the surgery does not enter the skull influence your view on invasiveness?
    5. Again, considering other diseases such as [Epilepsy, Depression, Rheumatoid Arthritis, Heart failure] does the invasiveness of VNS seem appropriate for the disease it would be treating?
4. BARRIERS
    1. Do you have any concerns about practical barriers to accessing these therapies, if patients want to use them? Are there things that you think might keep people from being able to use these treatments? (e.g., distance, waiting times, side effects, transportation, insurance, cost, access to information, caregiver support, work schedules, stigma)?
        1. Prompt as needed: Are you aware of the programming requirements for VNS and depression? Do you think it will be possible for you to attend your programming sessions, or would it be a barrier for you?
    2. What do you see as the most important ethical questions we should answer about using VNS for treating [depression/epilepsy/ OTHER] Why? How so?

    c. Do you think the barriers to VNS depend on the indication?

    d. When you think of the word ‘support’ in relation to VNS what comes to mind?
    Prompt: What would make people feel more supported?
    Prompt: Any specific concerns about demographics such as race, age, sex, culture, religion, etc.

***Interview Questions***

Patient (Epilepsy)

1. GENERAL
    1. Briefly, how long have you lived with Epilepsy? How many types of treatments have you tried?
    2. How does VNS compare to other therapies you have tried?

3. VNS is used for both epilepsy and depression. From your understanding of the disease, do you think VNS for epilepsy is more, less, or about the same level of benefit compared to depression?

2. RISK
    1. Is VNS more, less, or about the same risk level as [TREATMENT]
        i. Prompt as needed: slow onset of VNS
    1. Does VNS have more, less, or about the same amount of benefit as [TREATMENT]
        1. Prompt as needed: about expectations regarding medication reduction or potential for less side effects
    2. Does VNS have more, less, or about the same amount of side effects as [TREATMENT]
    3. Are the risks and side effects worth the potential benefits of VNS? What are the side effects expected from VNS? Compared to [TREATMENT] do you have any control over the side effects of VNS?
        1. Prompt as needed: about surgical complications, perceived inability to do certain imaging procedures
    4. If you had another disease such as rheumatoid arthritis or heart failure and were offered VNS as a treatment would you consider it? Why or why not? [WILLINGNESS]
3. INVASIVENESS
    1. How would you describe how invasive VNS is?  Why?  In what sense?
    2. Do you think VNS is more or less invasive than undergoing psychotherapy? Why? How so?
    3. Do you think VNS is more or less invasive than taking pharmaceuticals?  Why?  How so?
    4. Do you think VNS is more or less invasive than excision surgery? Why?  How so?
        1. Prompt as needed: Does the fact that the surgery does not enter the skull influence your view on invasiveness?
    5. Again, considering other diseases such as [Epilepsy, Depression, Rheumatoid Arthritis, Heart failure] does the invasiveness of VNS seem appropriate for the disease it would be treating?
4. BARRIERS
    1. Do you have any concerns about practical barriers to accessing these therapies, if patients want to use them? Are there things that you think might keep people from being able to use these treatments? (e.g., distance, waiting times, side effects, transportation, insurance, cost, access to information, caregiver support, work schedules, stigma)?
        1. Prompt as needed: Are you aware of the programming requirements for VNS and depression? Do you think it will be possible for you to attend your programming sessions, or would it be a barrier for you?
    2. What do you see as the most important ethical questions we should answer about using VNS for treating [depression/epilepsy/ OTHER]  Why?  How so?
    c. Do you think the barriers to VNS depend on the indication?
    d. When you think of the word ‘support’ in relation to VNS what comes to mind?
        Prompt: What would make people feel more supported?
        Any specific concerns about demographics such as race, age, sex, culture, religion, etc.

***Interview Questions***

Caregiver (Depression)

1. GENERAL
    1. Briefly, how long has your loved one lived with Depression?
    2. How did you learn about VNS? (Loved one, physician, internet search, etc.)
    3. What was your initial reaction to your loved one’s interest in VNS?
    4. VNS is used for both epilepsy and depression. From your understanding of the disease, do you think VNS for epilepsy is more, less, or about the same level of benefit compared to depression?
2. RISK
    1. Is VNS more, less, or about the same risk level as [TREATMENT]?
        i. Prompt as needed: slow onset of VNS
    1. Does VNS have more, less, or about the same amount of benefit as [TREATMENT]?
        1. Prompt as needed: about expectations regarding medication reduction or potential for less side effects
    2. Does VNS have more, less, or about the same amount of side effects as [TREATMENT]?
    3. Are the risks and side effects worth the potential benefits of VNS? What are the side effects expected from VNS? Compared to [TREATMENT] do you have any control over the side effects of VNS?
        1. Prompt as needed: about surgical complications, perceived inability to do certain imaging procedures
    4. If your loved one had another disease such as rheumatoid arthritis or heart failure and were offered VNS as a treatment would you want them to consider it? Why or why not?
3. INVASIVENESS
    1. How would you describe how invasive VNS is? Why? In what sense?
    2. Do you think VNS is more or less invasive than undergoing psychotherapy? Why? How so?
    3. Do you think VNS is more or less invasive than taking pharmaceuticals? Why? How so?
    4. Do you think VNS is more or less invasive than Deep Brain Stimulation? Why? How so?
        1. Prompt as needed: Does the fact that the surgery does not enter the skull influence your view on invasiveness?
    5. Again, considering other diseases such as [Epilepsy, Depression, Rheumatoid Arthritis, Heart failure] does the invasiveness of VNS seem appropriate for the disease it would be treating?
4. BARRIERS
    1. Do you have any concerns about practical barriers to accessing these therapies, if patients want to use them? Are there things that you think might keep people from being able to use these treatments? (e.g., distance, waiting times, side effects, transportation, insurance, cost, access to information, caregiver support, work schedules, stigma)?

        1. Prompt as needed: Are you aware of the programming requirements for VNS and depression? Do you think it will be possible for you to attend your programming sessions, or would it be a barrier for you?
    2. What do you see as the most important ethical questions we should answer about using VNS for treating [depression/epilepsy/ OTHER]  Why?  How so?
    c. Do you think the barriers to VNS depend on the indication?
    d. When you think of the word 'support' in relation to VNS what comes to mind?
        Prompt: What would make people feel more supported?
        Prompt: Any specific concerns about demographics such as race, age, sex, culture, religion, etc.

***Interview Questions***

Caregiver (Epilepsy)

1. GENERAL
    1. Briefly, how long has your loved one lived with Epilepsy?
    2. How did you learn about VNS? (Loved one, physician, internet search, etc.)
    3. What was your initial reaction to your loved one's interest in VNS?
    4. VNS is used for both epilepsy and depression. From your understanding of the disease, do you think VNS for epilepsy is more, less, or about the same level of benefit compared to depression?
2. RISK
    1. Is VNS more, less, or about the same risk level as [TREATMENT]?
        i. Prompt as needed: slow onset of VNS
    1. Does VNS have more, less, or about the same amount of benefit as [TREATMENT]?
        1. Prompt as needed: about expectations regarding medication reduction or potential for less side effects
    2. Does VNS have more, less, or about the same amount of side effects as [TREATMENT]?
    3. Are the risks and side effects worth the potential benefits of VNS? What are the side effects expected from VNS? Compared to [TREATMENT] do you have any control over the side effects of VNS?
        1. Prompt as needed: about surgical complications, perceived inability to do certain imaging procedures
    4. If your loved one had another disease such as rheumatoid arthritis or heart failure and were offered VNS as a treatment would you want them to consider it? Why or why not?
3. INVASIVENESS
    1. How would you describe how invasive VNS is?  Why?  In what sense?
    2. Do you think VNS is more or less invasive than undergoing diet changes? Why? How so?
    3. Do you think VNS is more or less invasive than taking pharmaceuticals?  Why?  How so?
    4. Do you think VNS is more or less invasive than excision surgery? Why?  How so?
        1. Prompt as needed: Does the fact that the surgery does not enter the skull influence your view on invasiveness?

5. Again, considering other diseases such as [Epilepsy, Depression, Rheumatoid Arthritis, Heart failure] does the invasiveness of VNS seem appropriate for the disease it would be treating?

4. BARRIERS
    1. Do you have any concerns about practical barriers to accessing these therapies, if patients want to use them? Are there things that you think might keep people from being able to use these treatments? (e.g., distance, waiting times, side effects, transportation, insurance, cost, access to information, caregiver support, work schedules, stigma)?
        1. Prompt as needed: Are you aware of the programming requirements for VNS and depression? Do you think it will be possible for you to attend your programming sessions, or would it be a barrier for you?
    2. What do you see as the most important ethical questions we should answer about using VNS for treating [depression/epilepsy/ OTHER] Why? How so?
    c. Do you think the barriers to VNS depend on the indication?
    d. When you think of the word 'support' in relation to VNS what comes to mind?
        1. Prompt: What would make people feel more supported?
        2. Prompt: Any specific concerns about demographics such as race, age, sex, culture, religion, etc.

**Surveys**

**Demographics Survey**

Please fill out the following demographic questions. These questions will help researchers understand the background of those who participate in this study.

- Birth Year
- What sex were you assigned at birth?
    - Male
    - Female
    - Other
- How do you currently describe yourself?
    - Male
    - Female
    - Transgender
    - I use a different term (open text)
- Which of the following do you consider yourself to be? Check all that apply
    - American Indian or Alaska Native
    - Asian
    - Black or African American
    - Hispanic, Latino or Spanish Origin
    - Middle Eastern or North African
    - Native Hawaiian or Other Pacific Islander
    - White
    - Other
    - Prefer not to disclose

- Roughly how much income from all sources (including earnings, pensions, investments, etc.) did your household have last year (before taxes)?
    - Less than $15,000
    - $15,000 to $24,999
    - $25,000 to $34,999
    - $35,000 to $49,999
    - $50,000 to $74,999
    - $75,000 to $99,999
    - $100,000 ot greater
    - Don’t know or prefer not to answer
- The dollar amount for direct payment influenced my decision to participate in REVEAL?
    - Strongly disagree, disagree,,neither agree or disagree, agree, strongly agree
- How would you describe the REVEAL study activities?
- Have you EVER been employed for wages or salary?
    - Yes
    - No
    - I don’t know
- If yes, what kind of work have you done most of your life? (What is/was your job called?)
    - 
    - Other
- What is your highest level of education?
    - Some high school
    - High school graduate
    - Some college
    - Trade/Technical/Vocational training
    - Some postgraduate work
    - Postgraduate degree
- What is your marital status?
    - Single, never married
    - Married or domestic partnership
    - Widowed
    - Divorced
    - Separated
- Including yourself, how many people currently live in your household?
    - 1, 2, 3, 4, or more than 4
- How many of those individuals are children?
    - #
- Which of the following diagnoses do you have? [Patient] / Which of the following diagnoses does your loved one have? [Caregiver] / Which disease do you work with regularly? [Provider]
    - Epilepsy
    - Depression
- What type of treatments have you tried for your disease? [Patient Only]
    - Medication

  - Therapy
  - Diet Change
  - Light Therapy
  - Transcranial Magnetic Stimulation
  - Electroconvulsive Therapy
  - Other (please describe)

**Knowledge Survey**

Please fill out the following VNS knowledge questions.

TRUE/FALSE Questions

- Knowledge (Epilepsy only)
  - VNS is negatively affected by cell phones
  - VNS is effective in treating epilepsy
  - VNS cannot be used with anti-epileptic medications
  - VNS is an outdated treatment
  - VNS can take up to 2 years to reach full effectiveness
  - VNS is typically painful when stimulation is on
  - VNS surgery is done while awake
  - VNS stops all seizures
  - VNS implants are MRI safe
  - VNS continuously sends stimulation
  - VNS is covered by insurance

- Knowledge (Depression only)
  - VNS is negatively affected by cell phones
  - VNS is effective in treating depression
  - VNS cannot be used with psychiatric medications
  - VNS is an outdated treatment
  - VNS can take up to 2 years to reach full effectiveness
  - VNS is typically painful when stimulation is on
  - VNS surgery is done while awake
  - VNS cures depression fully
  - VNS implants are MRI safe
  - VNS continuously sends stimulation
  - VNS is covered by insurance

Resource Questions

- How confident are you about your knowledge of VNS?
  - confident scale from ‘Not very confident’ to ‘Confident’
- Do you feel you have been given enough resources to learn about VNS?
  - Yes/No

**Attitudes Survey**

Please fill out the following questions. These questions will help researchers understand how people feel about VNS.

**Scale** = Extremely Unlikely, Somewhat Unlikely, Unlikely, Likely, Somewhat Likely, Extremely Likely

1. How likely is VNS to cause positive effects?
2. How likely is VNS to cause negative effects?
3. How likely are you to recommend VNS to someone who might need it?

**Scale** = Yes / No

4. Have you considered other neuromod options? (Y/N)
    1. If so, which ones?
5. Have you previously tried to pursue VNS? (Y/N) **[patients only]**
    1. If yes, did your insurance cover VNS?
    2. If yes, why did you not pursue VNS previously?

**Scale** = Doubtful, Somewhat Doubtful, Unsure, Somewhat hopeful, Hopeful

f. Fill in this sentence. I feel _______ that VNS as a treatment is an effective treatment for myself? **[Patients only]**

g. Fill in this sentence. I feel _______ that VNS as a treatment is an effective treatment for my loved one? **[Caregivers Only]**

h. Fill in this sentence. I feel _______ that VNS as a treatment is an effective treatment for my patients? **[Clinicians Only]**

**<u>Stigma Survey</u>**

Think about your experience living with [DISEASE] when filling out these questions.

Scale = 1 = Never, 2 = Rarely, 3 = Sometimes, 4 = Often, and 5 = Always

1. Because of my illness, some people seemed uncomfortable with me.
2. Because of my illness, some people avoided me.
3. Because of my illness, I felt left out of things
4. Because of my illness, people were unkind to me
5. Because of my illness, people avoided looking at me
6. I felt embarrassed about my illness
7. I felt embarrassed because of my physical limitations
8. Some people acted as though it was my fault I have this illness

Now, imagine that you have [ALT DISEASE]. Take a moment to think about how your life would be different. Complete the below questions as if you were living with [ALT DISEASE].

1. Because of my illness, some people seemed uncomfortable with me.
2. Because of my illness, some people avoided me.
3. Because of my illness, I felt left out of things
4. Because of my illness, people were unkind to me
5. Because of my illness, people avoided looking at me
6. I felt embarrassed about my illness

7. I felt embarrassed because of my physical limitations
8. Some people acted as though it was my fault I have this illness

**Rankings**

VNS is being explored for many types of diseases. We want to know more about how people feel this technology can be used in other diseases. There are no right or wrong answers.

Scale = Not appropriate, Somewhat appropriate, Very appropriate

How appropriate is VNS for treating Depression?
How appropriate is VNS for treating Epilepsy?
How appropriate is VNS for treating Rheumatoid Arthritis?
How appropriate is VNS for treating Heart Failure?

Please rank the following items from least to most invasive

| Depression | Epilepsy |
|---|---|
| Psychotherapy<br>TMS<br>ECT<br>VNS<br>DBS<br>Medication<br>Ketamine<br>Cingulotomy | Diet Changes<br>Medication<br>Excision Surgery<br>VNS<br>DBS |

**Barriers Survey**

People often face barriers to getting the care they need. Read through the following list and select items that you have experienced in getting care for your disease.

1. Distance to healthcare facility
2. Lack of knowledge
3. Stigma
4. Lack of family/social support
5. Denial/minimization of symptoms
6. Cost of treatment
7. Side effects of treatment
8. Inefficacy of treatment
9. Discrimination
10. Help-seeking is a sign of weakness
11. Afraid of what people might think if I sought help
12. Did not trust medical providers
13. Long wait times
14. Did not know who to contact to get help

From the items you selected, please rank their influence from least influential to most influential on your ability to get care for your disease.

**Support Activities Survey [Caregivers Only]**

In the last TWO YEARS, have you been the PRIMARY CAREGIVER for a family member, spouse, or close friend with [DISEASE]?

By “care,” we mean helping them with daily activities, driving them to appointments, offering emotional support, etc.

No, I have no family member, spouse, or close friend with [DISEASE]
No, I have a family member, spouse, or close friend with [DISEASE], but I did not provide care for them.
Yes, I have provided care for a family member, spouse, or close friend with [DISEASE].

How are you related to this person?
immediate family (parent, child, sibling)
extended family (cousin, grandparent, in-law)
spouse
close friend
neighbor, co-worker, or acquaintance

Over the last TWO YEARS, what kinds of support have you provided REGULARLY? Select all that apply.

- offering emotional support via talking, texting, and e-mailing
- running errands around town
- cleaning inside or outside of home
- preparing meals
- taking care of pets or companion animals
- providing financial support
- driving them to medical appointments
